# Refining cardiometabolic risk assessment using MRI-derived pancreas volume and fat content: insights from the NAKO and UK Biobank

**DOI:** 10.64898/2026.01.15.26344167

**Authors:** Matthias Jung, Zeynep Berkarda, Marco Reisert, Susanne Rospleszcz, Tobias Pischon, Thoralf Niendorf, Hans-Ulrich Kauczor, Henry Völzke, Katharina Laubner, Christopher L. Schlett, Michael T. Lu, Jochen Seufert, Fabian Bamberg, Vineet K. Raghu, Jakob Weiss

## Abstract

**Background:** The pancreas is essential for metabolic homeostasis. Alterations in morphology and parenchymal integrity may impact proper function but are not routinely used for risk stratification. Here, we propose an AI-pipeline to quantify pancreas volume and fat content from MRI to identify individuals at high-risk for cardiometabolic disease in the general population.

**Methods:** We quantified pancreas volume (milliliters, mL) and intrapancreatic fat content (defined as fat fraction; FF, %) from MRI of UK Biobank (UKB) and German National Cohort (NAKO) participants using deep learning. We 1) analyzed differences in volume and FF across age and sex, 2) computed percentile-curves and z-scores adjusted for age and sex to identify high-risk volumes/FF, and 3) conducted Cox regression to assess associations between z-score categories (volume: reference, z=−1 to 1; low, z=<−1; high, z>1; FF: low, z<1; moderate, z=0-1; high, z>1) and incident outcomes (diabetes, major adverse cardiovascular events (MACE), all-cause mortality) after adjustment for risk factors.

**Results:** Among 63,548 UKB and NAKO-participants (57.7±12.8 years; BMI: 26.3±4.4 kg/m^2^, 46.9% female), automated pancreas analysis revealed a positive association between both volume and FF and age. In 33,099 UKB-participants (median 4.8 years follow-up), z-score categories were associated with incident diabetes (low volume, aHR:1.59, 95%CI[1.20-2.11]; high FF, aHR:1.70, 95%CI[1.31-2.19]), MACE (high volume, aHR: 0.79, 95%CI[0.61-1.01]; high FF, aHR: 1.32, 95%CI[1.01-1.73]), and all-cause mortality (low volume, aHR: 1.48, 95%CI[1.16-1.90]) beyond risk factors. Adding z-score categories to a baseline model including risk factors improved discrimination of future diabetes (volume:0.781 to 0.784, p=0.004; FF:0.781 to 0.787, p<0.001) and mortality (volume:0.781 to 0.787, p<0.001)

**Conclusions:** Deviations from normalized pancreas volume and FF predicted cardiometabolic outcomes beyond known risk factors and alcohol intake. This automated approach identifies high-risk individuals who may benefit from cardiometabolic/endocrinology referral.

## INTRODUCTION

Cardiometabolic diseases, including cardiovascular disease and type 2 diabetes, were accountable for 19 million deaths in the United States between 1999 and 2020.^1^ Projections estimate that cardiovascular disease alone will cause 35.6 million deaths globally by 2050, making it the leading cause of death worldwide.^2^ The pancreas plays a central role in cardiometabolic disease development through its regulation of glucose metabolism and insulin secretion.^3,4^ Pancreatic dysfunction, including beta-cell failure and pancreatic fat content, contributes to insulin resistance and type 2 diabetes (T2D), key drivers of cardiovascular disease.^3,4^ Reduced volume and increased fat content of the pancreas are known to be associated with impaired glucose metabolism and a higher risk of T2D^5,6^ and could be measured from routine medical imaging studies like magnetic resonance imaging (MRI). However, this is currently not performed in daily practice due to the lack of reliable, automated postprocessing software and a limited number of studies investigating the clinical value of such measures beyond traditional clinical risk factors like BMI, hypertension and glucose testing.

Advances in artificial intelligence have made it possible to automatically and accurately measure the pancreatic volume and fat content from chemical shift encoding (CSE) MRI.^7,8^ As abdominal MRI is becoming an increasingly available and widely used medical imaging test, there is great potential for large-scale opportunistic screening, allowing for automated quantification of pancreas volume and fat content for risk assessment, regardless of the indication the scan was initially ordered for.^9,10^ A major challenge in utilizing pancreatic imaging features such as volume and fat content for risk assessment is the lack of population reference curves across different age decades, body size and sex as they are known to substantially influence pancreatic morphology and change over the lifespan.^11^

Here, we developed and tested a fully automated 3D deep learning (DL) framework to estimate pancreatic volume and fat fraction (FF) as a proxy for intra-pancreatic fat content from CSE MRI of over 63,000 individuals from the general population. We then described age- and sex-specific distributions of pancreatic volume and FF across the lifespan and calculated age-, sex-, and height-adjusted z-scores for pancreas volume and age- and sex-adjusted scores for FF. We assessed whether this novel z-score approach was associated with future T2D, major adverse cardiovascular events (MACE), and all-cause mortality beyond known cardiometabolic risk factors and alcohol intake. Finally, we released a free, web-based z-score calculator to accelerate future research and increase reproducibility between studies and populations.

## MATERIAL AND METHODS

### Overview of the study design

We propose a deep learning framework that automatically quantifies three-dimensional (3D) pancreas volume and FF from MRI. First, we visualized their distributions across the lifespan and calculated age- and sex-stratified percentile curves. Second, we investigated the association between pancreas volume and FF z-scores and future T2D, MACE, and all-cause mortality. Lastly, we developed a web-based calculator to enable the comparison of pancreas volume and FF against percentile curves from a large Western European population. An overview of the study design is provided in **Figure 1**.

**Figure 1:**
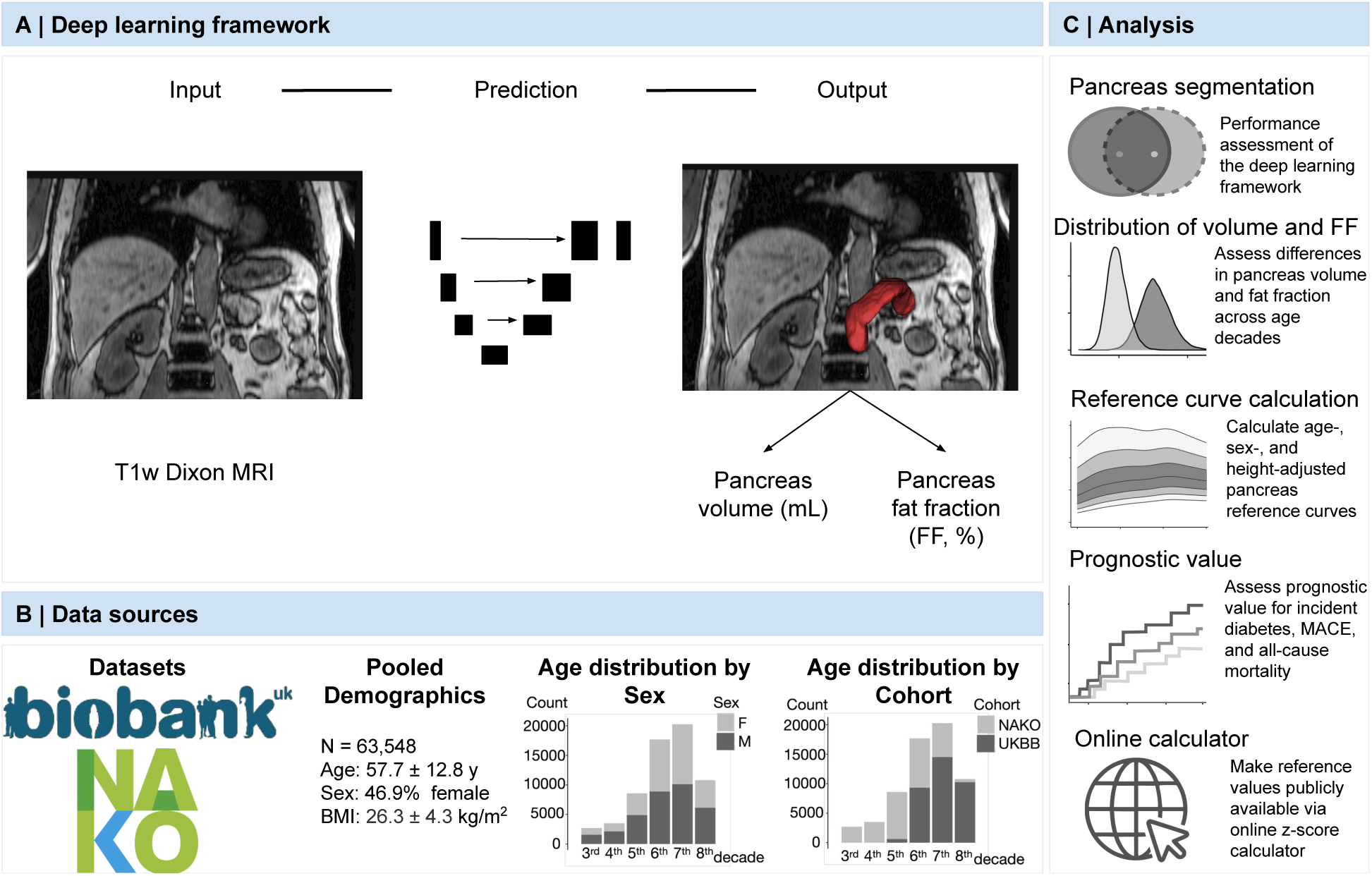
Overview of the study design. **(A)** The deep learning framework was developed to estimate pancreas volume (L) and fat fraction (FF, %) from MRI. **(B)** Pancreas segmentation was performed on whole-body MRIs in more than 63,000 individuals of two large population-based cohort studies, the UKB (36,314 individuals) and the NAKO (30,350 individuals). **(C)** After assessment of the pancreas segmentation, we investigated the differences in pancreas volume and FF distributions across age decades, calculated age-, sex-, (and height-) adjusted percentile curves and derived z-scores, investigated their prognostic value for incident health outcomes, and made available a web-based z-score calculator. Biobank, UK Biobank. FF, fat fraction. L, liters. MACE, major adverse cardiovascular event. MRI, magnetic resonance imaging. NAKO, German National Cohort. Y, years.

### Data sources

This study used data from two large population-based cohort studies: 1) the UK Biobank (UKB) and 2) the German National Cohort (NAKO).^12,13^

The UKB is an ongoing large, prospective population-based study investigating the genetic and non-genetic determinants of diseases of middle and older age. It included over 500,000 volunteers aged 38-73 years at baseline across the United Kingdom and provides detailed clinical parameters on prevalent diseases and outcomes.^12,14^

The NAKO is an ongoing interdisciplinary epidemiologic cohort study investigating disease prevention and prognosis, focusing on major disease groups such as cardiovascular disease, diabetes, and cancer, with 205,415 volunteers aged 19-74 years enrolled at 18 sites in Germany.^15^

Both studies included a comprehensive MRI examination in a subgroup of participants, which included a whole-body T1-weighted 3D VIBE two-point Dixon sequence. In this study, we used this sequence to quantify pancreas volume and FF. Further detailed information on the data sources is provided in **Supplemental Methods.**

### Deep learning framework development and testing

We developed and tested a fully automated deep learning framework to quantify pancreas volume and FF from T1-weighted 3D VIBE two-point Dixon MRI. The framework consisted of two models: 1) model 1 to segment the pancreas on whole-body T1-weighted Dixon MRI, and 2) model 2 that corrected for block-wise Dixon swaps in the UKB before FF extraction.

#### Development

For training, an experienced radiologist performed manual annotations of the pancreas using all four image contrasts of the T1-weighted Dixon MRI sequence (in-phase, opposed-phase, water, fat) and three imaging planes (axial, coronal, and sagittal reconstructions). These segmentations (n=150 randomly selected NAKO participants) were independently validated and adjusted where necessary by an attending board-certified radiologist. The inputs to the deep learning model were all four contrasts of the T1-weighted two-point VIBE Dixon MR sequence (in-phase, opposed-phase, fat image, and water image); the output of the model was a segmentation mask of the pancreas. In addition, the pancreas fat fraction (FF, %) was derived from the CSE-based water-fat information of the VIBE Dixon sequence as follows:

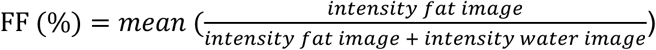

#### Testing

The model was independently tested in additional randomly selected NAKO (n=50 internal) and UKB (n=50 external) data sets. Annotations were generated in a similar manner as mentioned above. Model performance was assessed using the DICE similarity coefficient and Pearson’s correlation coefficient.

Further detailed information on model development and testing (also for model 2) is provided in **Supplemental Methods, Supplemental Results**, and **Supplemental Figure 3**.

### Distribution of Pancreas Volume and FF in pooled UKB and NAKO data

Age- and sex-stratified distributions of pancreatic volume and FF as well as age-, sex-, and height-adjusted reference percentile curves were determined through pooled analysis of all NAKO and UKB participants who had available pancreatic MRI data along with demographic information (age, sex, and height).

### Covariates for outcome analysis in the UKB only

The following a priori defined clinical covariates were considered: age, sex, BMI, waist circumference, race, alcohol consumption, smoking status, and hypertension in the UKB. In the NAKO, only age, sex and height were used. Detailed information on the extraction and definition of covariates from UKB and NAKO is provided in **Supplemental Methods.**

### Outcomes in the UKB only

All survival analyses were performed in the UKB only, as outcome data were not available for NAKO participants. Outcomes of interest were 1) incident T2D (ICD-10: E11; ICD-9: 250), 2) incident MACE (myocardial infarction: ICD-10: I21-22; ICD-9: 410-411; ischemic stroke: ICD-10: I63; ICD-9: 433-434; mortality from major cardiovascular diseases: ICD10: I1-6; I70-78) and 3) all-cause mortality. Outcomes were defined using UKB Data Fields 41270 and 41271 which contain distinct International Classification of Disease 9 and 10 diagnosis codes through linkage to all hospital inpatient records (https://biobank.ctsu.ox.ac.uk/crystal/refer.cgi?id=138483). All-cause mortality and mortality from major cardiovascular diseases was extracted through linkage to national death registries. Follow-up time in the UKB was calculated as the date of the MRI examination until the earliest date among death, outcome, loss to follow-up, October 31, 2022 for ICD-based outcomes (T2D and MACE), or May 25, 2023 (date of UKB data download) for all-cause mortality. “Prevalent” outcomes were diagnoses prior to the MRI, and “incident” outcomes were diagnoses after the MRI date.

### Statistical Analysis

#### Baseline characteristics

Baseline characteristics of the study participants are presented as mean ± standard deviation (SD) or median with interquartile ranges (IQR) for continuous variables and absolute counts with percentages for categorical variables.

#### Pancreas volume and FF across age decades and percentile curves

First, differences in pancreas volume and FF across age decades were visualized using density plots and quantified using the median and interquartile ranges (IQR). Then, sex-stratified percentile curves based on age and height (pancreas volume) or age only (FF) were created. We fit sex-stratified generalized additive models (GAMs) with isotropic smooth terms for age and height for pancreas volume to predict the conditional mean of pancreas volume using the mgcv R package (version 1.9-1, 2023, open source). A second GAM was developed using the same predictors to predict the conditional variance of pancreas volume. Because FF data was right-skewed, we fit sex-stratified quantile generalized additive models (qGAMs) with isotropic smooth terms for age to estimate the conditional percentiles (3rd, 10th, 25th, 50th, 75th, 90th, and 97th) of FF using the qgam R package (version 1.3.4, 2023, open source). Separate qGAMs were fit for each percentile. We performed internal 5-fold cross-validation to assess the median absolute deviation of both GAMs (**Supplemental Table 1**). The derivatives of the 50^th^ percentiles were calculated to illustrate the rate of change of pancreas volume and FF over the lifespan of an average-sized female (1.65 m) and male (1.75 m).

#### Association between age-, sex-, and height-specific z-scores and outcomes in the UKB

Outcome analyses were only performed in the UKB and limited to individuals without prevalent diabetes or a history of stroke or myocardial infarction. An individual’s z-score for pancreas volume and FF was calculated as follows:

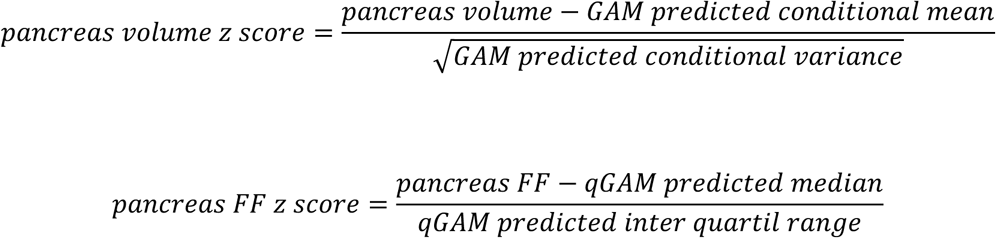

In addition to age and sex, pancreas volume z-score was adjusted for height rather than body mass index or body surface area, as height is predominantly genetically determined and less affected by modifiable factors such as diet and physical activity. The association between age-, sex-, and height-specific z-scores of pancreas volume and age- and sex-specific z-scores for FF and health outcomes was explored for z-score categories defined as “reference” (z-score = −1 to 1), “low” (z-score < −1) and “high” (z-score > 1) for pancreas volume and “reference” (z-score < 0), “moderate” (z-score = 0 to 1) and “high” (z-score > 1) for FF. Thus, an individual’s pancreas volume was classified as “high” if it was greater than one conditional-standard deviations above the age-, sex-, and height-specific conditional mean. Likewise, an individual’s pancreas FF was classified as “high” if it was greater than 1 conditional-interquartile range above the conditional-median based on age and sex.

To investigate time to outcome in the UKB, cumulative incidence curves (Kaplan-Meier method) and log-rank tests were computed using the above-defined z-score categories. The associations between the z-score categories and incident outcomes were evaluated via multivariable Cox proportional hazards regression analysis adjusted for the above-mentioned demographics and risk factors. These confounders were identified using a modified disjunctive cause criterion. To assess whether pancreas measures carry incremental value beyond these risk factors, nested Cox proportional hazards models with and without pancreas measures were compared using Harrell’s C-index and likelihood ratio test. The proportional hazards assumption was tested by computing scaled Schoenfeld residuals, and linearity was assessed using Martingale residuals. Both assumptions were satisfied for all models. Results from Cox regressions were reported as hazard ratio (HR) and 95% confidence intervals (CI). All statistical analyses were performed using R V4.2.2 (R Core Team, www.r-project.org, 2022).

## RESULTS

### Study population

The study population consisted of 63,548 individuals (n=34,697 UKB and n=28,851 NAKO participants; 29,832 (46.9%) females and 33,717 (53.1%) males with a mean age of 57.7±12.8 years and a mean BMI of 26.3±4.4 kg/m^2^. Pancreas volume (75.3±21.9 mL vs. 57.6±17.3 mL) and FF (9.6% (IQR 7.2-14.4%) vs. 7.4% (IQR 5.7-10.2%)) were significantly higher in males than females (both p<0.001; **Table 1**).

**Table 1:** Baseline characteristics – full study population.

| Characteristic | Overall, N = 63,548<sup>1</sup> | Female, N = 29,832<sup>1</sup> | Male, N = 33,716<sup>1</sup> |
| --- | --- | --- | --- |
| Age (y) | 57.7 ± 12.8 | 58.1 ± 12.2 | 57.3 ± 13.4 |
| BMI (kg/m<sup>2</sup>) | 26.3 ± 4.4 | 25.9 ± 4.8 | 26.7 ± 3.9 |
| Pancreas volume (mL) | 67.0 ± 21.8 | 57.6 ± 17.3 | 75.3 ± 21.9 |
| Pancreas FF (%) | 8.5 (6.4-12.4) | 7.4 (5.7-10.2) | 9.6 (7.2-14.4) |
| **Data Source** |  |  |  |
| NAKO | 28,851/63,548 (45%) | 12,320/29,832 (41%) | 16,531/33,716 (49%) |
| UKBB | 34,697/63,548 (55%) | 17,512/29,832 (59%) | 17,185/33,716 (51%) |

<sup>1</sup> Mean ± SD; Median (IQR); n/N (%)

### Differences in pancreas volume and FF across age decades

First, we analyzed the differences in median pancreas volume and FF and their IQR across age decades stratified by sex (**Figure 2A**). Median pancreas volume increased until the 7^th^ decade and then slightly decreased in both sexes (**Figure 2A**). In both sexes, the variability of pancreas volume increased until the 6^th^ decade and then slightly decreased. We observed a positive association between FF and age, with higher variability in males than females that increased across the lifespan (from 4.6% (IQR 3.8-5.5%) in the 3^rd^ decade to 9.0% (IQR 6.9-12.6%) in the 8^th^ decade in females and from 5.6% (IQR 4.5-7.1%) to 11.3% (IQR 8.3-16.7%) in males; **Figure 2B**).

**Figure 2:**
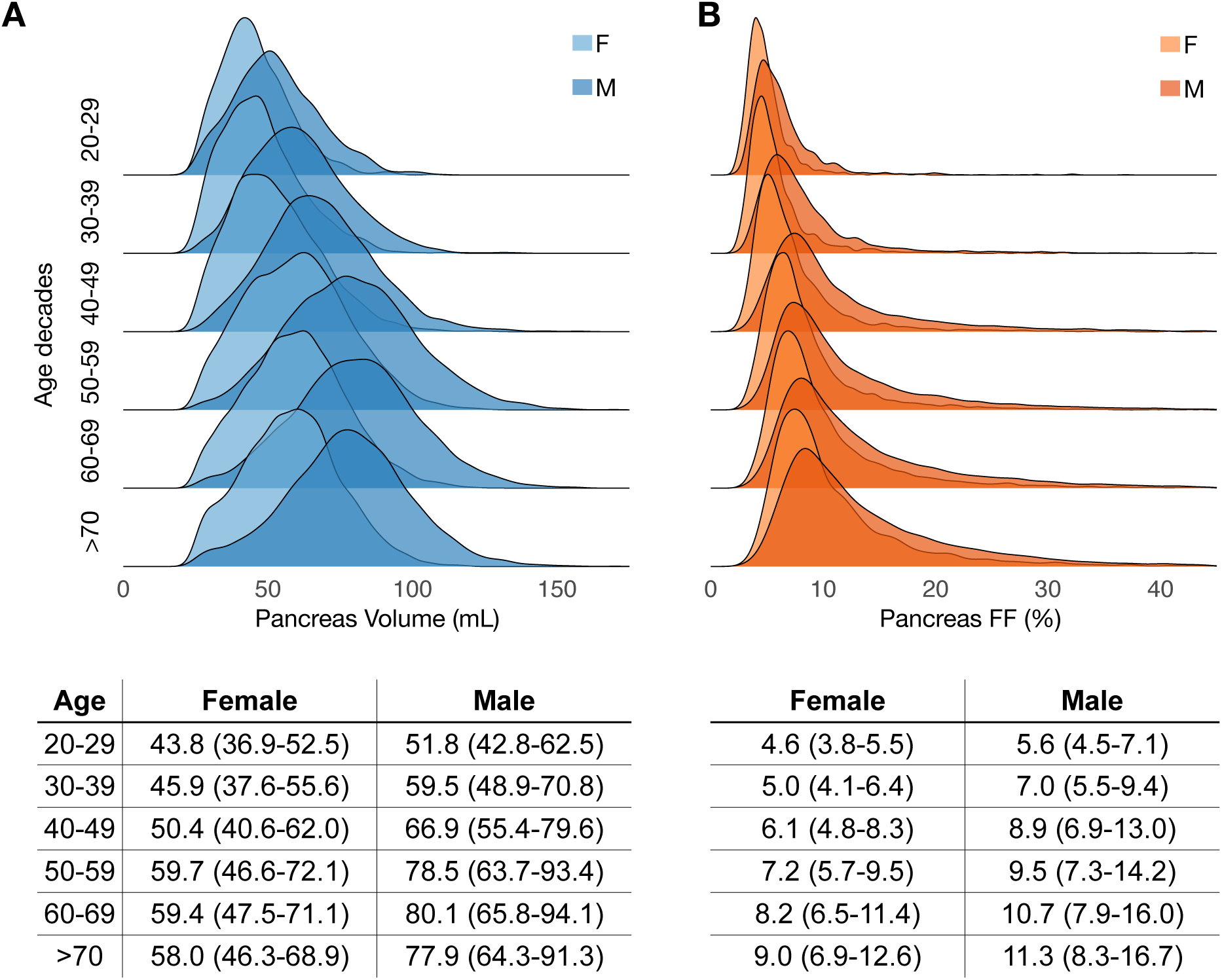
Pancreas volume and FF distribution across age decades. Density plots illustrate the differences in pancreas volume (blue), and FF (orange) across age decades. While median pancreas volume **(A)** increased until the age of 60 and then slightly decreased in both sexes, median FF **(B)** increased throughout the lifespan in both sexes, with higher variability in males than females. Sex-stratified median (IQR) pancreas volume and FF are provided in the tables below the plots. F, female. FF, fat fraction. IQR, interquartile range. L, liters. M, male.

### Age, sex, and height-adjusted pancreas volume and FF percentile curves

Next, we calculated age-, sex-, and height-specific percentile curves for pancreas volume and FF (**Figure 3**). Crude values for the pancreas volume and FF across age stratified by sex are shown as a scatter plot in **Figure 3A**. Percentile curves were calculated using a Generalized Additive Model (GAM) fit stratified by sex using smooth functions of age (FF) and age and height (pancreas volume; see **Methods**). Examples of the resulting percentile curves are shown for an average-height female (1.65 m tall; **Figure 3B**) and an average-height male (1.75 m tall; **Figure 3C**). The variance in pancreas volume and FF was higher in males than in females across all ages (**Figure 3B & 3C**).

**Figure 3:**
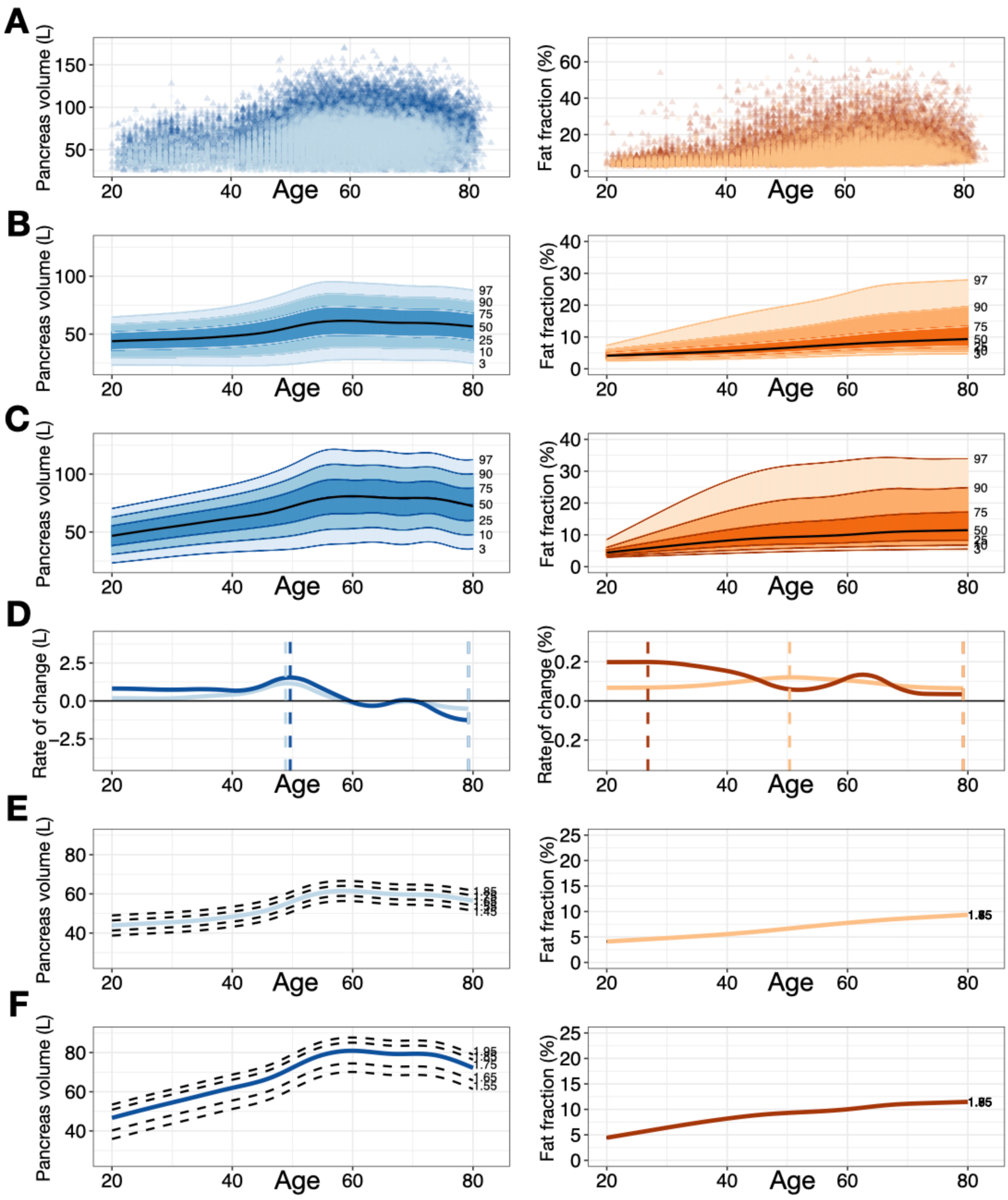
Percentile curves for pancreas volume and fat fraction. **(A)** Scatterplots of crude pancreas volume and FF as a function of age (stratified by sex). **(B & C)** Graphs show age-, sex-, and height-adjusted pancreas volume percentile curves with 3^rd^, 10^th^, 25^th^, 50^th^, 75^th^, 90^th^, and 97^th^ percentile lines for a 1.65 m tall female **(B)** and a 1.75 m tall male **(C)**. Pancreas volume variance was higher in average males than in average females across all ages. **(D)** The derivatives of the 50^th^ percentiles were calculated to illustrate the rate of change of pancreas volume over the lifespan of an average 1.65 m tall female and 1.75 m tall male. Dashed lines indicate the minimum and maximum rate of change. **(E & F)** Graphs show the 50^th^ percentile of females **(E)** and males **(F)** of different body heights (female: 1.45-1.85 m; male: 1.55-1.85 m). Pancreas volume showed a height-related dependency in both sexes **(E & F).** F, female. L, liters. M, male.

Pancreas volume increased in both sexes until 7^th^ decade after which it began to decrease (**Figure 3D**). Pancreas FF increased throughout the lifespan in both sexes (**Figure 3D**). Pancreas volume showed a height-related dependency in both sexes (**Figure 3E & 3F).**

### Association between pancreas volume and FF z-scores and health outcomes

To investigate whether deviations from the typical age-, sex-, and height-related changes established in the percentile curve analyses predict incident health outcomes, we next examined the association between pancreas volume and FF z-score categories with incident T2D, MACE, and all-cause mortality in the UKB (no outcome data was available for NAKO participants). The final cohort consisted of 33,099 individuals (17,054 females and 16,045 males) with a mean age of 64.7±7.8 years and a mean BMI of 25.9±4.1 kg/m^2^ after excluding individuals with prevalent T2D or a history of MACE before the date of their MRI examination (**Table 2**). Over a median follow-up of 4.2 years (IQR 3.3-5.6 years), 503/33,099 (1.5%) individuals were diagnosed with T2D, 530/33,099 (1.6%) individuals experienced MACE, and 525/33,099 (1.6%; median follow-up 4.8 years [IQR 3.9-6.2]) individuals died of any cause (**Table 2**).

**Table 2:** Cohort characteristics - UKB testing cohort for evaluation of association between pancreas variables and outcomes.

| Characteristic | Overall, N = 33,099 <sup>1</sup> | Female, N = 17,054<sup>1</sup> | Male, N = 16,045<sup>1</sup> |
| --- | --- | --- | --- |
| Age (y) | 64.7 ± 7.8 | 64.1 ± 7.6 | 65.4 ± 7.9 |
| BMI (kg/m<sup>2</sup>) | 25.9 ± 4.1 | 25.5 ± 4.4 | 26.3 ± 3.7 |
| **Fat Fraction** |  |  |  |
| Category |  |  |  |
| z<0 | 18,633/33,099 (56%) | 9,263/17,054 (54%) | 9,370/16,045 (58%) |
| z=0-1 | 10,177/33,099 (31%) | 5,514/17,054 (32%) | 4,663/16,045 (29%) |
| z>1 | 4,289/33,099 (13%) | 2,277/17,054 (13%) | 2,012/16,045 (13%) |
| **Pancreas Volume** |  |  |  |
| Category |  |  |  |
| z<-1 | 22,778/33,099 (69%) | 11,462/17,054 (67%) | 11,316/16,045 (71%) |
| z=-1-1 | 3,629/33,099 (11%) | 2,188/17,054 (13%) | 1,441/16,045 (9.0%) |
| z>1 | 6,692/33,099 (20%) | 3,404/17,054 (20%) | 3,288/16,045 (20%) |
| Race (White) | 32,057/33,016 (97%) | 16,522/17,018 (97%) | 15,535/15,998 (97%) |
| **Weekly Alcohol** |  |  |  |
| Consumption | 23,744/32,850 (72%) | 11,190/16,914 (66%) | 12,554/15,936 (79%) |
| **History of** |  |  |  |
| Hypertension | 4,262/33,099 (13%) | 1,801/17,054 (11%) | 2,461/16,045 (15%) |
| Ever smoked | 13,353/32,953 (41%) | 6,370/16,968 (38%) | 6,983/15,985 (44%) |
| Incident Diabetes | 503/33,099 (1.5%) | 163/17,054 (1.0%) | 340/16,045 (2.1%) |
| **Incident MACE** | 530/33,099 (1.6%) | 163/17,054 (1.0%) | 367/16,045 (2.3%) |
| Follow up time (y): | 4.2 (3.4-5.6) | 4.2 (3.4-5.6) | 4.2 (3.3-5.6) |
| T2D and MACE |  |  |  |
| **All-cause Death** | 525/33,099 (1.6%) | 185/17,054 (1.1%) | 340/16,045 (2.1%) |
| **Follow up time (y):** | 4.8 (3.9-6.2) | 4.8 (3.9-6.2) | 4.8 (3.9-6.2) |
| **Death** |  |  |  |

<sup>†</sup> Mean ± SD; n/N (%); Median (IQR)

Cumulative incidence curves showed graded associations between pancreas volume and FF z-score categories for all investigated outcomes (**Figures 4 & 5**).

**Figure 4:**
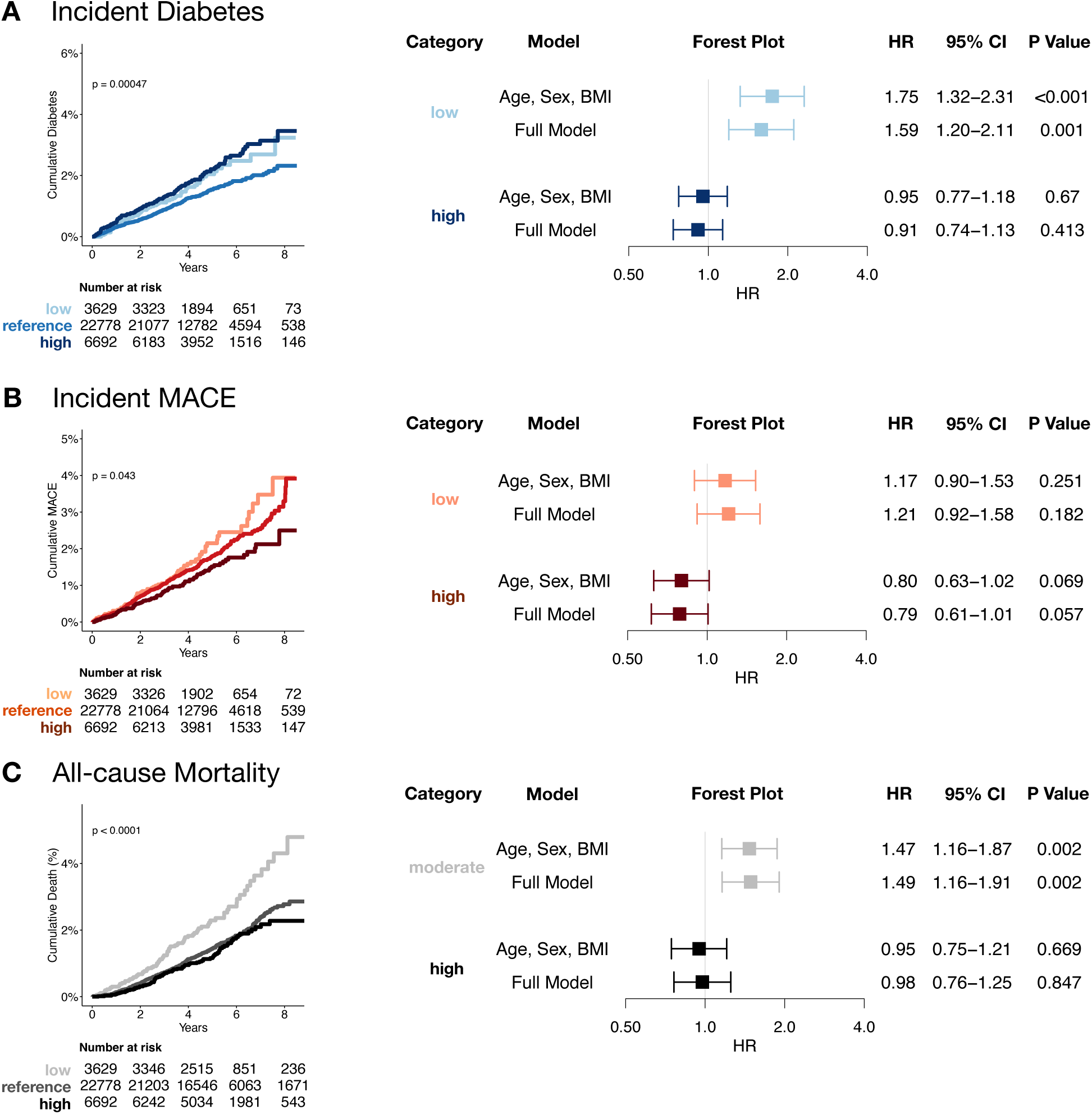
Cumulative incidence curves and forest plots for pancreas volume z-score categories and incident diabetes, MACE and all-cause mortality in the UKB. (**A**) Incident diabetes, (**B**) MACE, and (**C**) all-cause mortality in the UKB after excluding individuals with prevalent diabetes and a history of MACE (**Table 2**) according to pancreas volume z-score categories (reference, z<0; moderate, z=0-2; high, z>2). Cumulative incidence and Kaplan-Meier curves show significant differences between the pancreas volume z-score categories. Forest plots show hazard ratios with 95% confidence intervals of Cox proportional hazard regression analyses. Models were adjusted for 1) age, sex, and BMI, and 2) additional adjustment for traditional risk factors including waist circumference, race, alcohol consumption, smoking status, and hypertension (full model). BMI, body mass index; CI, confidence interval; aHR, adjusted hazard ratio; MACE, major adverse cardiovascular events; UKB, UK Biobank;

**Figure 5:**
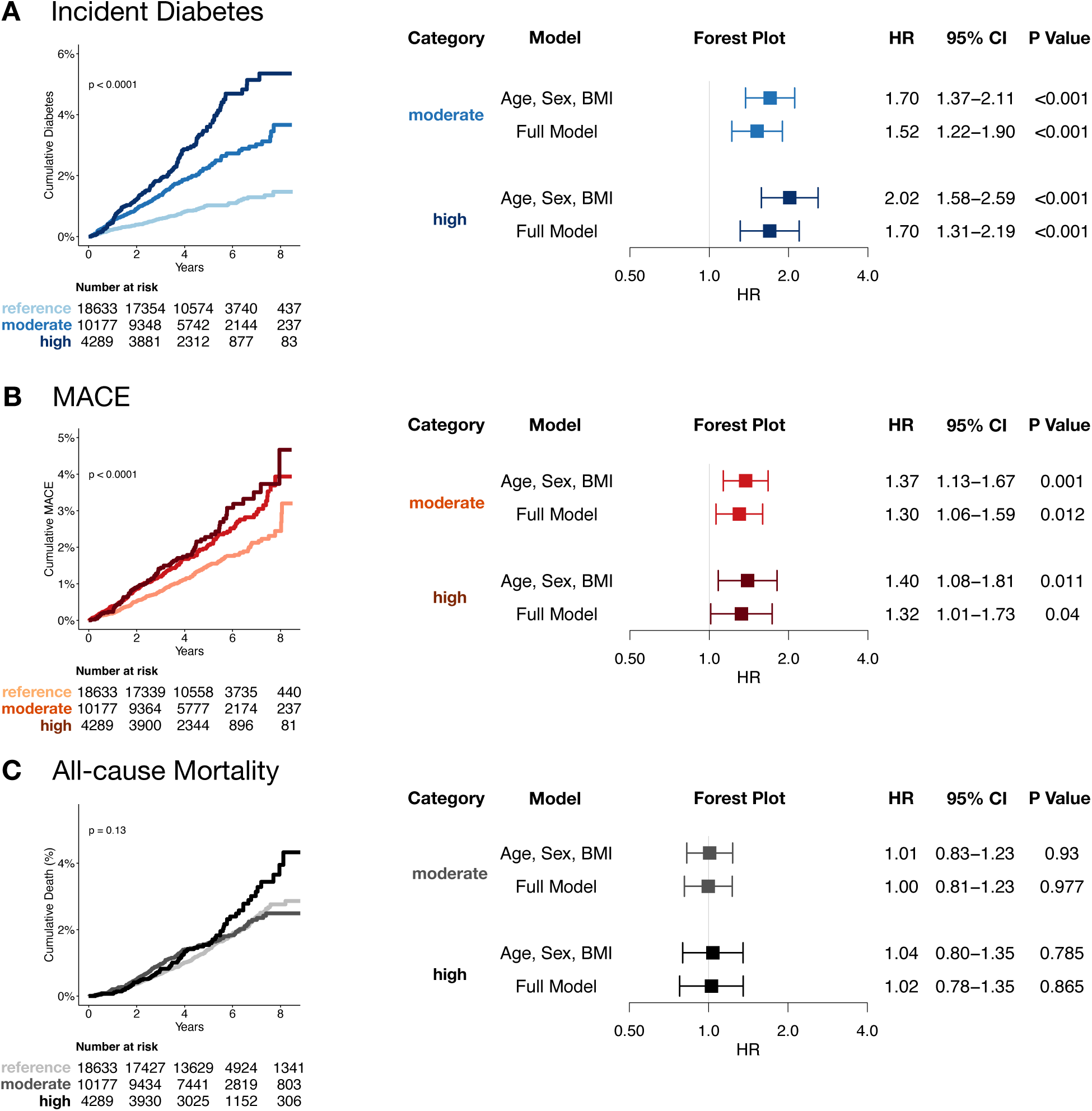
Cumulative incidence curves and forest plots for FF z-score categories and incident diabetes, MACE and all-cause mortality in the UKB. (**A**) Incident diabetes, (**B**) MACE, and (**C**) all-cause mortality in the UKB after excluding individuals with prevalent diabetes and a history of MACE (**Table 2**) according to FF z-score categories (reference, z<0; moderate, z=0-2; high, z>2). Cumulative incidence and Kaplan-Meier curves show significant differences between the pancreas volume z-score categories. Forest plots show hazard ratios with 95% confidence intervals of Cox proportional hazard regression analyses. Models were adjusted for 1) age, sex, and BMI, and 2) additional adjustment for traditional risk factors including waist circumference, race, alcohol consumption, smoking status, and hypertension (full model). BMI, body mass index; CI, confidence interval; FF, fat fraction; aHR, adjusted hazard ratio; MACE, major adverse cardiovascular events; UKB, UK Biobank;

#### T2D

Low pancreas volume z-scores were associated with higher incident T2D risk after adjustment for traditional clinical risk factors (age, sex, BMI categories, waist circumference, race, alcohol consumption, smoking status, hypertension) (aHR: 1.59, 95% CI [1.20-2.11]; **Figure 4A**). Similarly, both moderate (aHR: 1.52, 95% CI [1.22-1.90]) and high pancreas FF z-scores (aHR: 1.70, 95% CI [1.31-2.19]) were associated with an increased risk of incident T2D after adjustment for same risk factors (**Figure 5A**).

#### MACE

High pancreas z-volume category (aHR: 0.79, 95% CI [0.61-1.01]) was associated with a reduced risk for future MACE independent of the clinical risk factors (**Figure 4C**). In contrast, moderate and high FF z-score categories were associated with 30% and 32% risk increase of incident MACE after adjusting for above mentioned covariates (moderate, aHR: 1.30, 95% CI [1.06-1.59]; high, aHR: 1.32, 95% CI [1.01-1.73]; **Figure 5C**).

#### All-cause mortality

Low pancreas volume z-scores were associated with an increased risk of all-cause mortality independent of age, sex, BMI, race, alcohol consumption, smoking status, and hypertension (aHR: 1.48, 95% CI [1.16-1.90]) (**Figure 4B**). No association was found for pancreas FF z-scores.

### Incremental value of pancreas volume and FF z-scores

#### Volume z-score categories

Adding the pancreas volume to a baseline model, which included clinical risk factors only, resulted in a small but significant improvement in the discrimination of incident diabetes (C-index 0.781 to 0.784, p=0.004) and all-cause mortality (C-index 0.720 to 0.723, p=0.007).

#### FF z-score categories

Adding the pancreas FF to a baseline model of clinical risk factors only resulted in a modest improvement in the discrimination of incident diabetes (C-index 0.781 to 0.787, p<0.001) only.

## DISCUSSION

In this study, we developed a fully automated deep learning framework to quantify pancreas volume and FF from Dixon MRI to calculate age-, sex- and height-adjusted pancreas volume and age- and sex-adjusted FF percentile curves and to investigate whether deviations from typical pancreas volume and FF (z-scores) were associated with health outcomes beyond cardiometabolic risk factors in a large Western European population. Our main findings were that 1) pancreas volume increased until 60 years of age, then decreased in both sexes, while pancreas FF increased throughout lifetime, and 2) volume and FF z-score categories predicted health outcomes in the general population beyond traditional risk factors: The low volume category was associated with an 1.5-fold higher risk of future diabetes and a 1.5-fold increased risk of all-cause mortality, while the high volume category was associated with a 22% lower MACE risk. High pancreas FF was associated with a 1.7-fold higher risk of future diabetes and a 1.4-fold higher risk of MACE. In addition, pancreas volume and FF z-scores added incremental value to traditional risk factors. We envision these z-scores as a new risk marker to opportunistically identify individuals at high risk for cardiometabolic disease beyond established clinical pathways. To facilitate translation and comparability of pancreas volume and fat fraction z-scores, we developed a publicly available z-score calculator (https://circ-ml.github.io/) to allow research and clinicians benchmark their data against the values of a large Western European cohort.

Intra-pancreatic fat content is a known predictor of metabolic syndrome, T2D, and cardiovascular disease.^16–19^ Excess intra-pancreatic fat may impair β-cell function through several pathways, including lipotoxicity, oxidative stress, low-grade inflammation, and mitochondrial dysfunction within islet cells, ultimately reducing insulin secretion capacity.^20–22^ These effects appear to be at least partially independent of generalized adiposity.^23,24^ In addition to its diabetogenic effects, intra-pancreatic fat has also been associated with adverse cardiovascular risk profiles^25^ such as dyslipidemia, systemic inflammation, and subclinical atherosclerosis^19,26,27^ In addition, higher pancreatic fat content has been associated with coronary artery calcification.^28^ Besides intra-pancreatic fat content, pancreatic volume has also been associated with metabolic health.^6,29^ Previous studies have shown that a smaller pancreas is linked to reduced insulin secretion and an increased risk of type 2 diabetes.^6,29^ Furthermore, Martin et al. provided genetic evidence from Mendelian randomization analyses suggesting that decreased pancreas volume may cause type 2 diabetes.^6^ Here, we introduce a non-invasive way to identify high-risk individuals based on abnormal pancreatic volume and fat content from MRI. We find that this identifies individuals at high-risk independent of prevalent cardiometabolic risk factors or body habitus. In particular, 13% of our cohort had severe intra-pancreatic fat content, which was associated with a 1.7-fold higher risk of future diabetes, and a 1.3-fold increased risk of future MACE, after adjustment for cardiometabolic risk factors. Furthermore, 11% of our cohort had an abnormally low pancreas volume, which was associated with a 1.6-fold higher risk for diabetes and a 1.5-fold higher risk for all-cause mortality, respectively. Intriguingly, high pancreatic volume had a protective effect. It reduced the risk of future MACE by 21%.

Despite the high potential value of estimation pancreatic volume and FF from MRI, it is so far not performed in clinical radiology workflows due to time and postprocessing software constraints. Thanks to advances in artificial intelligence, fully automated quantification of such measures has now become feasible on a population scale. Our deep learning framework used in the present study was developed on a chemical shift encoded T1-weighted two-point Dixon MR sequence, which is widely used in daily abdominal MRI examinations.^30^ Hence, pancreas volume and FF can be automatically extracted from routine clinical Dixon-MRI scans, irrespective of their initial indication, facilitating integration into electronic medical records (EMR) without disrupting clinical and radiology workflows.

However, as there is only limited data available so far, there are currently no established cutoff values to define typical age, sex and height-adjusted ranges of pancreatic volume and fat content that identify individuals at risk for future cardiometabolic disease. For FF for example, multiple highly varying thresholds have been suggested, ranging from 3.9% to 10.4%.^21,22,31,32^ The results from this study address this knowledge gap by providing reference curves for pancreas volume and FF from over 63,000 Western Europeans of the general population. Along with the derived percentile curves and the free online tool released with this publication, we aim to support researchers and clinicians to compare their study cohort or patient’s against to those of the general population. Our approach has the potential to 1) improve the comparability and reproducibility of research studies, 2) facilitate clinical use by identifying “high-risk” individuals based on their pancreas volume and FF, and 3) enhance prevention strategies by identifying patients who could benefit from cardiometabolic or endocrinology referrals.

This study has limitations. First, the study population consists primarily of white Western European adults aged 20 years and older from the United Kingdom and Germany. Generalizability of our percentile curves to other racial/ethnic groups, children, and/or clinical patient data may be limited. Second, Dixon fat-water swapping artifacts are common calculation errors in clinical routine, meaning that a fat-only signal can be erroneously displayed in parts of the image where a water-only signal would be expected, resulting in incorrect Dixon-derived fat fraction calculations.^33,34^ While we corrected these errors in our study cohort before FF extraction, this may be a minor barrier to translating the Dixon-based fat fraction quantification approach into clinical practice. Third, we did not include pancreas enzymes in the analyses as they were collected eight years before imaging in the UKB and had a high proportion of missing values. Last, we only evaluated our framework in the general population. How and whether translation to clinical scenarios could impact patient care and clinical decision-making needs to be investigated in future studies.

In conclusion, we found that a deep learning framework robustly extracted pancreas volume and FF from Dixon MRI to calculate percentile curves and z-scores that allowed for identifying individuals at risk for incident cardiometabolic disease and mortality in the general population beyond known risk factors. We released a free online tool that allows researchers and clinicians to calculate pancreas volume and FF z-scores on their own data sets to improve comparability and reproducibility. Future studies will test whether this framework can use routine imaging in clinical care to opportunistically identify high-risk individuals and guide patient management.

## ACKNOWLEDGMENTS

This project was conducted using data from the German National Cohort (NAKO) (www.nako.de). The NAKO is funded by the Federal Ministry of Education and Research (BMBF) [project funding reference numbers: 01ER1301A/B/C, 01ER1511D, and 01ER1801A/B/C/D], federal states of Germany, and the Helmholtz Association, the participating universities and the institutes of the Leibniz Association. This research has been conducted using the UK Biobank Resource under Application Number 80337. We thank all participants who took part in the NAKO and UKB study and the staff of these research initiatives. MJ was funded by the Deutsche Forschungsgemeinschaft (DFG, German Research Foundation) - 518480401. VKR was funded by Norn Group Longevity Impetus Grant, NHLBI K01HL168231, and AHA Career Development Award 935176.

## DATA AVAILABILITY

Due to the restrictions imposed by the NAKO ethics committee, the data are not publicly available but can be requested from https://transfer.nako.de/transfer/index. The data from the UKB can be downloaded upon request from https://www.ukbiobank.ac.uk. The open-source online tool for z-score and percentile calculation will be available from https://circ-ml.github.io/.

## CODE AVAILABILITY

Code will be made available via bitbucket.

## ETHICS APPROVAL

Informed consent was obtained from all participants in the UK Biobank and the German National Cohort study. In addition, we received local IRB approval (IRB of the University of Freiburg: 23-1316-S1-retro and 24-1099-S1-retro).

## AUTHOR CONTRIBUTIONS

**Matthias Jung:** conceptualisation, data curation, formal analysis, investigation, methodology, project administration, resources, software, supervision, validation, visualisation, writing – original draft

**Zeynep Berkarda:** conceptualisation, data curation, formal analysis, investigation, methodology, project administration, resources, software, supervision, validation, visualisation, writing – original draft

**Marco Reisert:** conceptualization, data curation, formal analysis, methodology, resources, software, supervision, validation, writing – review & editing.

**Susanne Rospleszcz:** conceptualisation, data curation, formal analysis, investigation, methodology, project administration, resources, supervision, validation, writing – review & editing.

**Tobias Pischon:** conceptualisation, data curation, funding acquisition, methodology, project administration, resources, software, supervision, validation, writing – original draft, and writing – review & editing.

**Thoralf Niendorf:** conceptualisation, data curation, funding acquisition, methodology, project administration, resources, software, supervision, validation, writing – review & editing.

**Hans-Ulrich Kauczor:** conceptualisation, data curation, funding acquisition, methodology, project administration, resources, software, supervision, validation, writing – review & editing.

**Henry Völzke:** conceptualisation, data curation, funding acquisition, methodology, project administration, resources, software, supervision, validation, writing – review & editing.

**Katharina Laubner:** conceptualisation, data curation, funding acquisition, methodology, project administration, resources, software, supervision, validation, writing – review & editing.

**Christopher L. Schlett:** conceptualisation, data curation, funding acquisition, methodology, project administration, resources, software, supervision, validation, writing – review & editing.

**Michael T. Lu:** conceptualisation, data curation, investigation, methodology, project administration, resources, software, supervision, validation, writing – review & editing.

**Jochen Seufert:** conceptualisation, data curation, funding acquisition, methodology, project administration, resources, software, supervision, validation, writing – review & editing.

**Fabian Bamberg:** conceptualisation, data curation, funding acquisition, methodology, project administration, resources, software, supervision, validation, writing – review & editing.

**Vineet K. Raghu:** conceptualisation, data curation, formal analysis, investigation, methodology, project administration, resources, software, supervision, validation, visualisation, writing – original draft

**Jakob Weiss:** conceptualisation, data curation, formal analysis, investigation, methodology, project administration, resources, software, supervision, validation, visualisation, writing – original draft

