## Supplemental Materials for "Refining cardiometabolic risk assessment using MRI-derived pancreas volume and fat content: insights from the NAKO and UK Biobank"

### I SUPPLEMENTAL METHODS

#### Data sources

*UKB*

The UKB is a large, prospective population-based study established to investigate the genetic and non-genetic determinants of diseases of middle and older age. It included over 500,000 volunteers aged 38–73 years at baseline across the United Kingdom and provides detailed clinical parameters on prevalent diseases and outcomes.[1,2]

In this study, we included all UKB participants from the multimodal-imaging cohort who underwent $T_1$-weighted 3D two-point VIBE Dixon MRI in axial orientation (slice thickness of 4.5 mm, voxel size of 2.23 x 2.23 mm, matrix of 224 x 168 x 44 , repetition time [TR] of 6.69 ms, echo time [TE] of 2.39 and 4.77 ms; 1.5T MAGNETOM Aera, Siemens Healthineers, Erlangen, Germany) until the date of data download, May 25, 2023.[3] A total of 36,471 baseline MRIs were identified. 159 individuals were excluded due to corrupt or incomplete images, resulting in 36,312 individuals with available MRIs for pancreas quantification (**Supplemental Figure 1**).

*NAKO*

The NAKO is an ongoing interdisciplinary epidemiologic cohort study investigating disease prevention and prognosis, focusing on major disease groups such as cardiovascular disease, diabetes, and cancer, with 205,415 participants aged 19-74 years enrolled at 18 sites in Germany.[3] Of these, 30,861 participants underwent whole-body MRI (3T MAGNETOM Skyra, Siemens Healthineers, Erlangen, Germany) at five imaging sites in the imaging substudy including a whole-body $T_1$-weighted 3D two-point VIBE Dixon sequence in axial orientation (slice thickness of 3 mm, voxel size of 1.41 x 1.41 mm, matrix of 320 x 260 x 96, TR of 4.36 ms, TE of 1.23 and 2.46 ms). The NAKO was approved by all local institutional review boards of the five imaging sites, and written informed consent was obtained from all participants before enrollment.[4]

For the current study, we used MR images and clinical information from the second release that included 30,770 participants who underwent MRI between May 27, 2014 and September 30, 2019. A total of 418 individuals were excluded because of corrupt or incomplete images, resulting in 30,352 individuals with available whole-body MRIs for pancreas quantification (**Supplemental Figure 2**).

**Development of model 1- pancreas segmentation model**

The pancreas segmentation model was developed to quantify pancreas volume in liters (mL) from $T_1$-weighted two-point VIBE Dixon MRI. For all annotations, image-based calculations, model development, and testing, the open-source NORA - medical imaging platform (www.nora-imaging.org, Freiburg, Germany) was used. The model was developed using MRI data from a random sample of n=150 NAKO individuals.[4] To ensure the most accurate discrimination and segmentation of different tissues, an experienced radiology resident performed manual annotations of the pancreas using all four image contrasts of the $T_1$-weighted Dixon sequence (in-phase, opposed-phase, water, fat) and three imaging planes (axial, as well as coronal and sagittal reconstructions). These segmentations were independently validated and adjusted where necessary by a board-certified radiologist. The inputs to the deep learning model were all four contrasts of the $T_1$-weighted two-point VIBE Dixon MR sequence (in-phase, opposed-phase, fat image, and water image); the output of the model was a segmentation mask of the pancreas. We applied a three-dimensional morphological erosion to the original binary mask using a spherical structuring element with a radius of two voxels. This operation removes a two-voxel-thick layer from all surfaces of the mask and uniformly shrinks the segmented volume. The erosion step ensures that the additionally derived pancreas fat fraction (FF, %) values are derived from pancreatic gland tissue only and are not influenced by surrounding peripancreatic fat. Pancreas FF was derived from the chemical shift encoding-based water-fat information of the VIBE Dixon sequence as follows:

$$\mathrm{FF} = mean\ (\frac{intensity\ fat\ image}{intensity\ fat\ image + intensity\ water\ image})$$

The model was implemented as a hierarchical, patch-based stack of convolutional neural networks (CNN). The patchwork approach[5] uses nested patches of a fixed matrix size that decrease in physical size. A U-Net-type architecture is utilized in each scale, with the U-Net matrix size set to 32x32x32 voxels for all scales. A scale pyramid with a depth of four is employed. These settings were chosen based on the available hardware capacity, which limited sample size and pyramid size. The pyramid size was chosen to achieve a coarsest layer field-of-view of 200 x 200 x 400 mm and a final resolution of 2 x 2 x 4 mm. Intermediate levels of the pyramid were exponentially interpolated. The input to the network consists of concatenated fat, water, in- and out-of-phase contrasts. The architecture of the U-Net used is similar to the default UNet configuration[6], with feature dimensions (8,16,16,32,64) and max-pooling and transposed convolutions in the encoding and decoding layers, respectively. Each U-Net has n+8 output channels, with the first n=1 corresponding to the labels and used for intermediate loss computations. The total logits of n+8 outputs are passed to the next scale. The network is trained with the Adam optimizer[7] with a learning rate of 0.001 and using ten million patches for training of the network with a batch size of 32. The training took approximately four days. No systematic tuning was performed, and all labels were trained using binary cross-entropy per channel. More details about the model architecture are reported elsewhere.[5]

**Independent testing of model 1 – pancreas segmentation model**

First, the model was internally tested in an independent dataset of n=50 randomly chosen NAKO participants not seen during any part of model development. To evaluate model performance, automatically generated volumetric segmentations were compared with manual reference segmentations using the Dice coefficient to quantify spatial overlap. In addition, Pearson's correlation coefficients were calculated to assess the linear agreement between quantitative measures (volume and FF) derived from the manual and predicted segmentation masks.

Second, we used transfer learning to apply the model to UKBB participants and account for differences in imaging data. Fine tuning was performed using a random sample of n=200 UKBB participants, which were labeled as described above. Independent testing of the retrained UKBB model was performed on an additional random sample of n=50 subjects from the UKBB not seen during any part of model retraining. Model performance was assessed by comparing the automatically generated volumetric segmentations of the retrained model with the manual reference segmentations using the Dice coefficient and Pearson's correlation coefficient.

**Model 2 - Correction of fat-water swap artifacts in UKB Dixon MRI**

We used our previously developed and tested model to correct for Dixon swaps, which were present as block-wise swaps of the acquired slabs for stitching in the UKB[8]. In brief, we trained a network that predicts the sign of the opposed-phase contrast and the ground truth was generated from n=100 UKB scans that did not show any Dixon swaps. The inputs to the network were the in- and opposed-phase images. To correct for Dixon swaps, fat and water images were computed based on the predicted sign and compared to the original (scanner-side) computed images. When differences were higher than 100 signal intensity units became too large, the contrast was exchanged from fat to water or vice versa in the original images to correct for the former Dixon swap artifact. All corrections were performed on a slice level.

**Covariates**

*UKB*

Date of birth, sex, and race were extracted from the self-reported UKB baseline population characteristics. Due to the small number of non-white participants in the UKB imaging study, race was dichotomized into white and others. Physical measurements, including BMI (weight/height$^2$; kg/m$^2$), were obtained at the imaging visit. BMI categories were defined as < 25, 25-30, and ≥ 30. Alcohol intake, smoking status, and history of cancer were self-reported via a touchscreen questionnaire at the imaging visit. Alcohol intake was defined as “regular” (“Daily or almost daily”, “Three or four times a week”, “Once or twice a week”) and “occasional” (“One to three times a month”, “Special occasions only”, “Never”). Smoking was dichotomized by subsuming former and current smokers into “ever smokers”. Prevalent hypertension was defined as ICD-10 codes I10-15 or ICD-9 codes 401-405.

*NAKO*

Only baseline demographic data were available for the NAKO participants. This included age at MRI, sex, height (m), and weight (kg), which were assessed with standardized measuring devices at the imaging centers (all Stadiometer 274 for height and medical Body Composition Analyzer 515 for weight, both seca GmbH, Hamburg, Germany).

### II SUPPLEMENTAL RESULTS

**Independent testing of model 1**

Internal testing was performed in 50 randomly selected individuals of the NAKO. The mean dice coefficient, which compares the voxel-wise agreement between the model's predicted pancreas segmentation and the manual pancreas segmentation, was 0.88 ± 0.02. Pearson's correlation coefficients assessing the linear relationship between manual and automatically generated segmentation mask volumes and FF percentages were R=0.88 and R=0.99 for pancreas volume and FF (p values <0.001; **Supplemental Figure 3A**), respectively.

External testing was performed on 50 randomly selected participants of the UKB imaging sub-study. Mean Dice coefficient was 0.84±0.03 in the UKB. Pearson's correlation coefficients comparing the manual and automatically generated segmentation mask volumes and FF percentages were R=0.80 and R=0.98 (p values <0.001), respectively (**Supplemental Figure 3B**).

#### III SUPPLEMENTAL Tables

**Table S1: Median absolute deviation of pancreas volume and FF GAMs**

| | MAD | |
|---|---|---|
| **Sex** | **Pancreas volume (mL)** | **Pancreas FF (%)** |
| Female | 11.6 | 1.84 |
| Male | 13.5 | 2.95 |

FF, fat fraction. GAM, generalized additive model. mL, milliliters

#### IV SUPPLEMENTAL FIGURES

**Supplemental Figure 1: Flowchart Diagram-UKB**

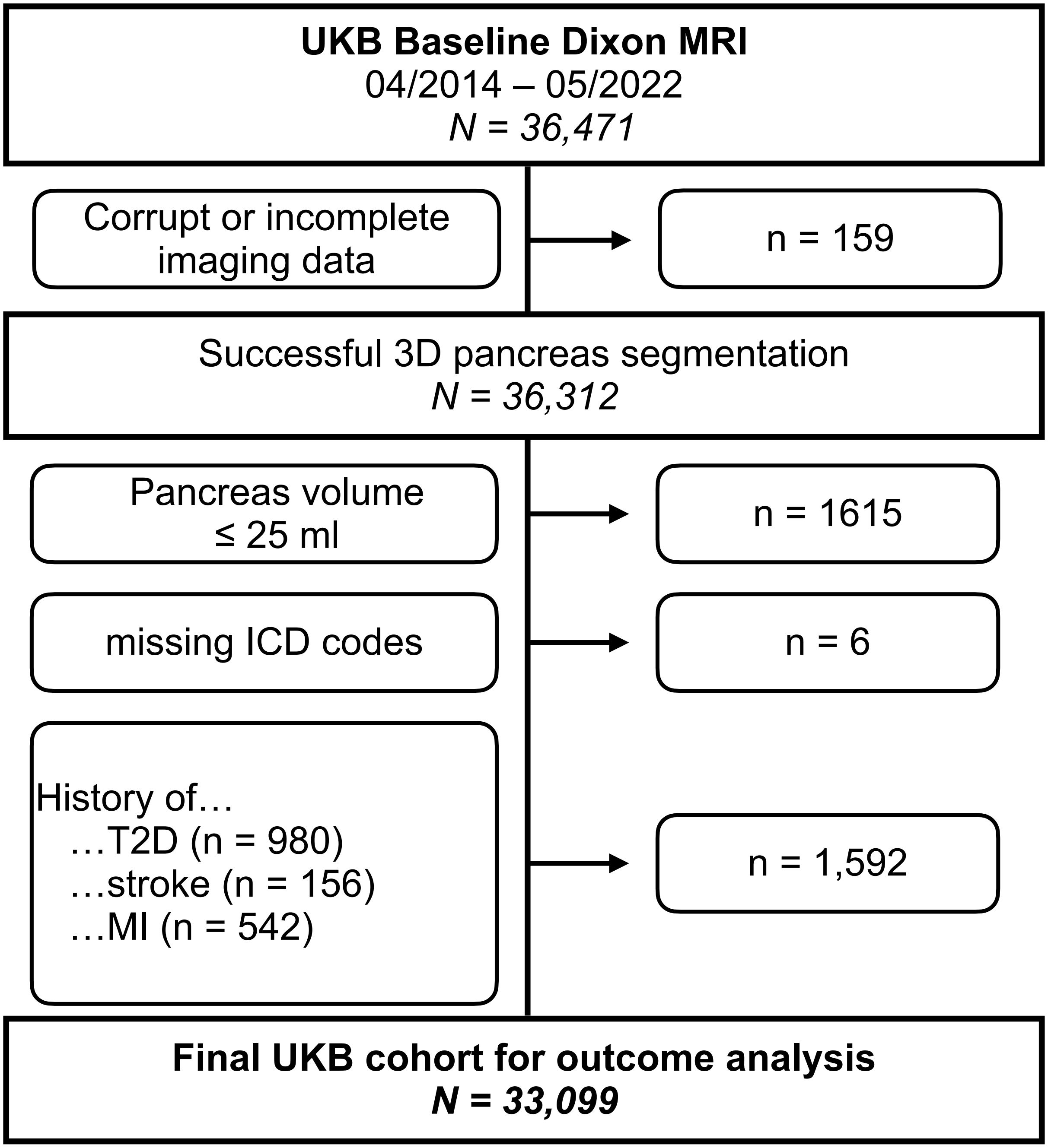

MI, myocardial infarction; MRI, magnetic resonance imaging. T2D, type 2 diabetes; UKB, UK Biobank;

**Supplemental Figure 2: Flowchart Diagram-NAKO**

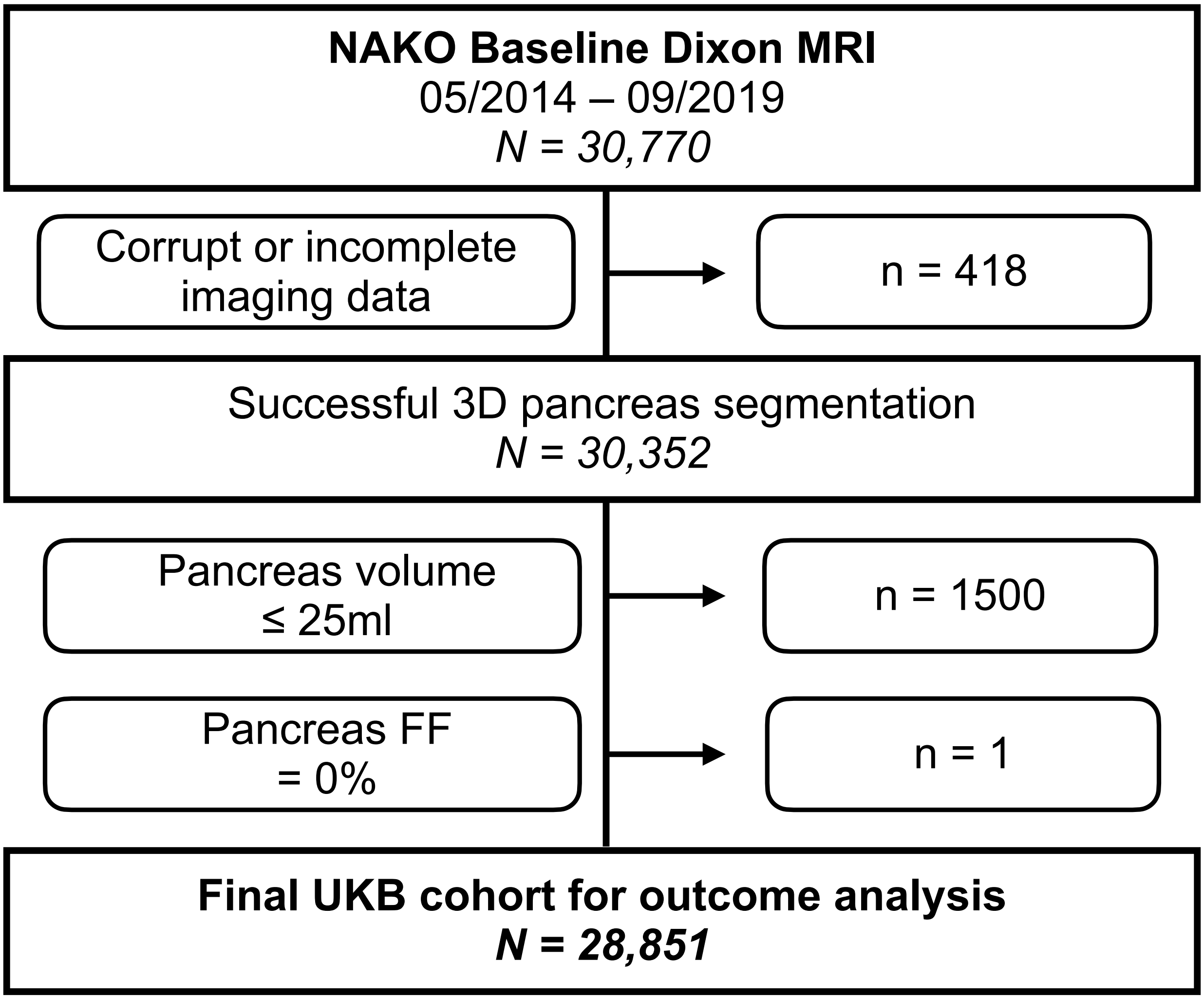

FF, fat fraction; MRI, magnetic resonance imaging. NAKO, German National Cohort.

**Supplemental Figure 3: Correlation between manual and deep learning-based 3D pancreas segmentations**

#### A NAKO – Independent testing dataset (n=50)

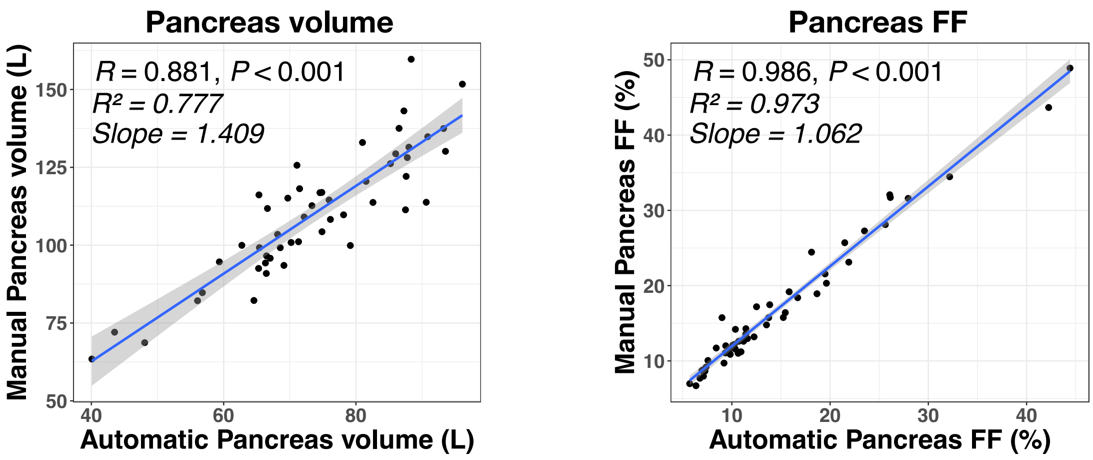

#### B UKBB – Independent testing dataset (n=50)

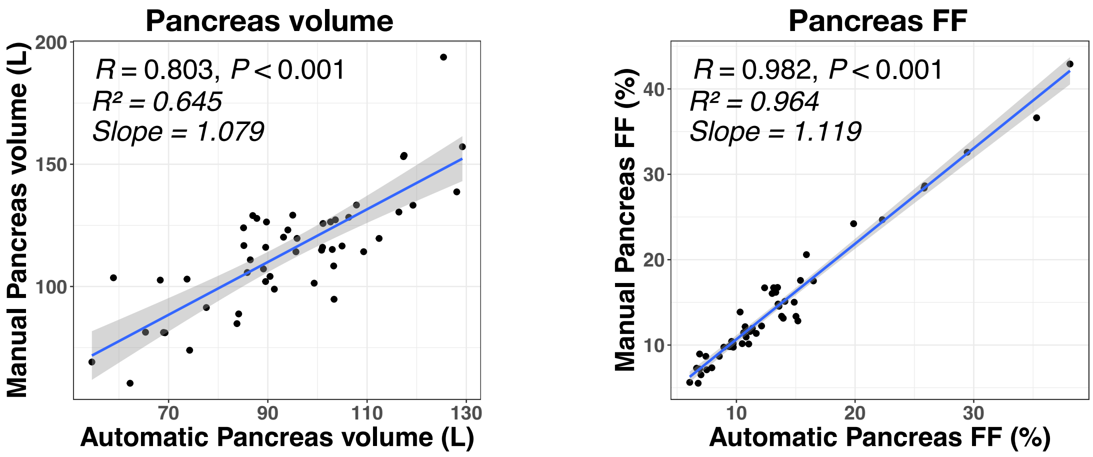

**Supplemental Figure 3 |** (**A**) Correlation between manual and deep learning-based 3D pancreas segmentations in the independent internal NAKO testing dataset (n=50) and (**B**) in the independent external UKB testing dataset (n=50) after transferring and fine-tuning on n=130 UKB datasets.

DL, deep learning. NAKO, German National Cohort. FF, fat fraction. UKB, UK Biobank.
